# The Somalia Mortality Estimation Database (S-MED): a Bird’s Eye View of Mortality and its Determinants

**DOI:** 10.1101/2025.09.07.25335291

**Authors:** Ruwan Ratnayake, Yamna Ouchtar, Yahye Abukar Ahmed, Jamal Hassan Mohamoud, Mohammed Jelle, Andrew Seal, Najib Isse Dirie, Jennifer Palmer, Francesco Checchi

## Abstract

Globally, there is a lack of consolidation and thus sharing of critical mortality estimates which can serve as an early warning of the severity of a humanitarian crisis. This lack of a comprehensive view may mask critical situations, and impact on which crises are more measured and visible to the humanitarian community. This ultimately affects the allocation of scarce humanitarian resources.

Somalia is a country marked by recurrent drought, armed conflict, food insecurity and malnutrition. To facilitate real-time investigation of mortality rates, we developed the open-source, publicly available Somalia Mortality Estimation Database (S-MED) to visualise mortality estimates from retrospective surveys and surveillance systems in Somalia in real-time. This enables improved awareness through visualization of mortality estimates as well as the capacity to facilitate multisectoral analysis of determinants of mortality (i.e., drought, displacement, disease outbreaks) and higher-level analysis (i.e., crisis-wide analysis of mortality estimates and mortality forecasting).

In this paper, we describe the mortality, morbidity, food insecurity, and environmental data contained in S-MED. We show how its mortality data can be used for improved detection of early warning signals and a more comprehensive public health interpretation of drought and armed conflict-driven health crises in 2018 and 2022. Similar mortality surveillance initiatives could be adapted to crisis-affected settings globally.

## Introduction

The most severe consequences of conflict and other natural disasters on health and survival are manifested as excess mortality, beyond the level that would be expected for the crisis-affected population.[1] In 2024, half of all deaths among children under 5 years of age occurred in 39 fragile and conflict-affected countries, including Niger, Nigeria, Somalia, Chad, and South Sudan.[2] Excess deaths are due to a cascade of distal and proximal risk factors including widespread civil conflict which negates investment in health systems; poor and unequal access to life-saving interventions including vaccination, the treatment of major childhood diseases, reproductive health care and safe childbirth; and underlying and persistent food insecurity and malnutrition.[1]

Civil registration and vital statistics systems in crisis-affected settings globally may be poorly functional, and typically do not provide adequate coverage for displaced populations.[1] Estimates of mortality in crisis-affected populations, while not meant to register deaths systematically, fulfil several goals including the estimation of the magnitude of mortality rates, forensic investigation into causes of death, and orienting resource allocation toward populations with the greatest need for humanitarian assistance.[1] Mortality estimates are typically computed as crude death rates (CDR) and under 5 year death rates (U5DR) and measured at a relatively small geographic scale (i.e., districts, displacement camps) using (a) retrospective mortality surveys (based on methods outlined for combined nutrition and mortality surveys by the Standardized Mortality and Assessment of Relief and Transitions (SMART) guidelines[3]) and (b) prospective mortality surveillance [4]. The geographical coverage and frequency of these small-scale efforts is by no means extensive or adequate among crisis-affected populations in a given country. Mortality and nutrition survey results are typically shared amongst partners of a country’s Health and Nutrition Clusters to guide and scale-up the response for a sudden onset or rapidly deteriorating situation.[5] However, achieving external visibility to the wider humanitarian system remains challenging, given that no centralized global repository of mortality estimates currently exists, and timely access to data in crises remains challenging.[6] A lack of processes for data sharing limits the urgent and meaningful use of these critical data across countries and often prolongs the masking of precarious situations of high and preventable mortality, as recently seen in particularly violent situations in Gaza, Central African Republic, Sudan and Eastern Democratic Republic of the Congo.[6] This influences which crises are more ‘measured’ and thus visible to the humanitarian community, with knock-on effects on the allocation of scarce humanitarian resources.

In the context of major reductions in global humanitarian and development funding, mortality data could be better used to guide resource allocation for the most severe of humanitarian crises. Global and/or crisis-wide repositories of mortality estimates, and associated health and risk factor data would vastly improve the visibility and investigation of mortality. Several prototypes have served different purposes including the Centre for Research on the Epidemiology of Disaster’s (CRED) Complex Emergency Database (CE-DAT) which was a global repository of mortality and nutrition surveys, but was concluded in 2012.[7] The Integrated Food Security Phase Classification (IPC, https://www.ipcinfo.org/) system maps and integrates food security and nutrition data in over 30 countries to enable classification of sub-national areas by the severity of acute malnutrition and acute and chronic food insecurity (as precursors of mortality). Within Somalia, the Food Security Nutrition and Analysis Unit’s (FSNAU) Early Warning-Early Action Dashboard (https://dashboard.fsnau.org/) aggregates predictor variables (i.e., displacement, insecurity, malnutrition admissions, food prices, etc.) to support projection of food insecurity. These efforts have in the past highlighted the situation in Somalia, where decades of armed conflict, underinvestment in health systems, drought, food insecurity, and health emergencies have resulted in consistently high child and crude mortality rates [8–10]. For instance, a simple mapping and plotting of mortality estimates after intense conflict in 2006 up to 2009 from surveys from Somalia contained in the CE-DAT database corroborated a marked increase in above-threshold CDR that were concentrated in southern Somalia.[8] Current food insecurity projections by the IPC take into account the anticipated impact of large-scale reductions in humanitarian aid funding, impending drought and reduced seasonal rainfall, high food prices, armed conflict and displacement influx, projecting that 36% of the population will experience acute food security (Phases 3 to 5) and further worsening of acute malnutrition across rural, riverine, pastoral, and agropastoral livelihood zones by June 2025.[11]

New approaches to centralise mortality estimates at the level of a given crisis would be beneficial for humanitarians to visualise and document mortality estimates in real-time (as well as to detect geographic gaps in coverage), share data in a common repository, and could facilitate crisis-wide estimation of mortality. There is also great value in interpreting the public health impacts of drought, violence, displacement and food insecurity by collating, visualizing, and analyzing predictor and mortality data in real-time to illustrate the interrelationships between morbidity and other determinants of mortality.[12] To help address these current gaps, we developed a data sharing and visualization platform called the Somalia Mortality Estimation Database (S-MED, https://hhc-lshtm.shinyapps.io/S-MED/). S-MED is driven by a visual interface to foster visibility, mapping, visualisation and ultimately public health interpretation of mortality estimates, and is underpinned by a repository of mortality estimates to facilitate crisis-wide level analysis of mortality in Somalia. In this article, we introduce S-MED, and present case studies of its use to support better interpretability of mortality estimates from Somalia.

## Methods

In this paper, we describe the objectives, design and analysis of S-MED, including case studies of its use during the 2017-2018 and 2022-2023 drought and armed conflict crises.

### Objectives and design

From the outset, we identified the following objectives for S-MED:

- Provide an open source and flexible platform to facilitate access to mortality data in Somalia
- Act as a repository to exhaustively include mortality estimates from organizations collecting mortality data in Somalia
- Allow users to visualise and map mortality estimates and time-trends to facilitate public health interpretation
- Develop situation analyses to benefit organizations collecting mortality and health data
- Support researchers to carry out higher-level analyses of crisis-wide mortality estimates

We discussed these objectives with public health and nutrition practitioners in Somalia (namely, UNICEF Somalia and Evidence for Change (e4c)) and from the SMART Initiative, experts at a mortality estimation consultative meeting at the US Centers for Disease Control and Prevention (CDC), and former CRED staff who managed the CE-DAT database. Their recommendations included to provide a visually-appealing design for non-epidemiologists and routine users of mortality, nutrition, and health data; to collate retrospective mortality estimates but also facilitate the prediction of mortality rates; to facilitate visualization and spatial analyses; to exhaustively integrate of mortality estimates from agencies collecting mortality data (i.e., NGOs, UN agencies, Ministry of Health, etc.) using retrospective surveys and prospective surveillance; and to use open source and secure data platforms that can be easily expanded or modified for other crises. The current design is outlined in Figure 1.

**Figure 1.**
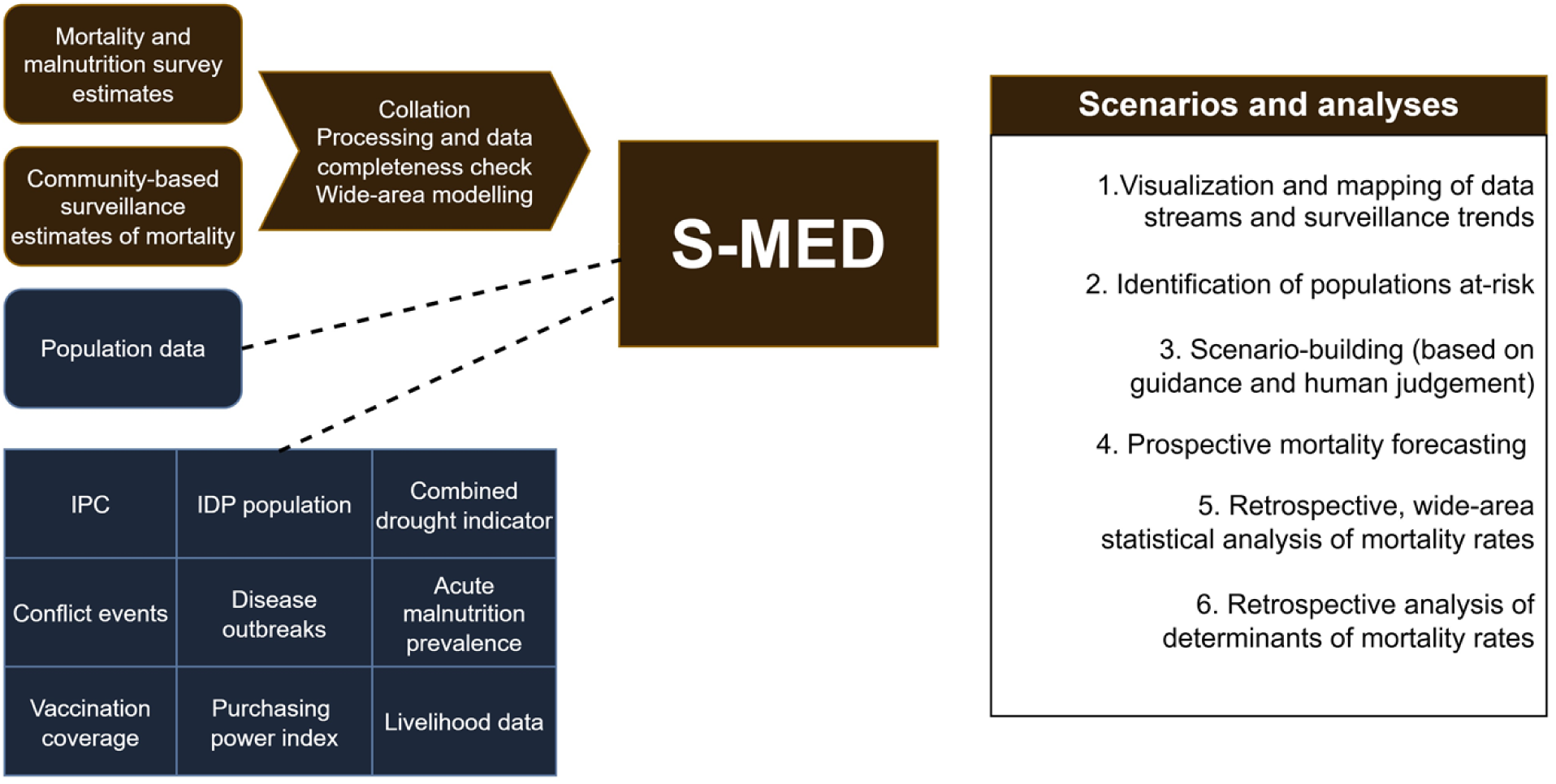
Framework for the Somalia Mortality Estimation Database (S-MED). IDP, internally displaced persons; IPC, Integrated Food Security Phase Classification.

We used R statistical software [13] and the Shiny framework with Bootstrap (https://shiny.posit.co/) to build a user-friendly interface hosted on the Shiny server and we copied the R code to a repository stored on GitHub (https://github.com/ruwanepi/S_MED) (Figure 2). Keeping the system open source is critical for encouraging the humanitarian community to adapt, remix and port this code for expansion of this project to other countries.

**Figure 2.**
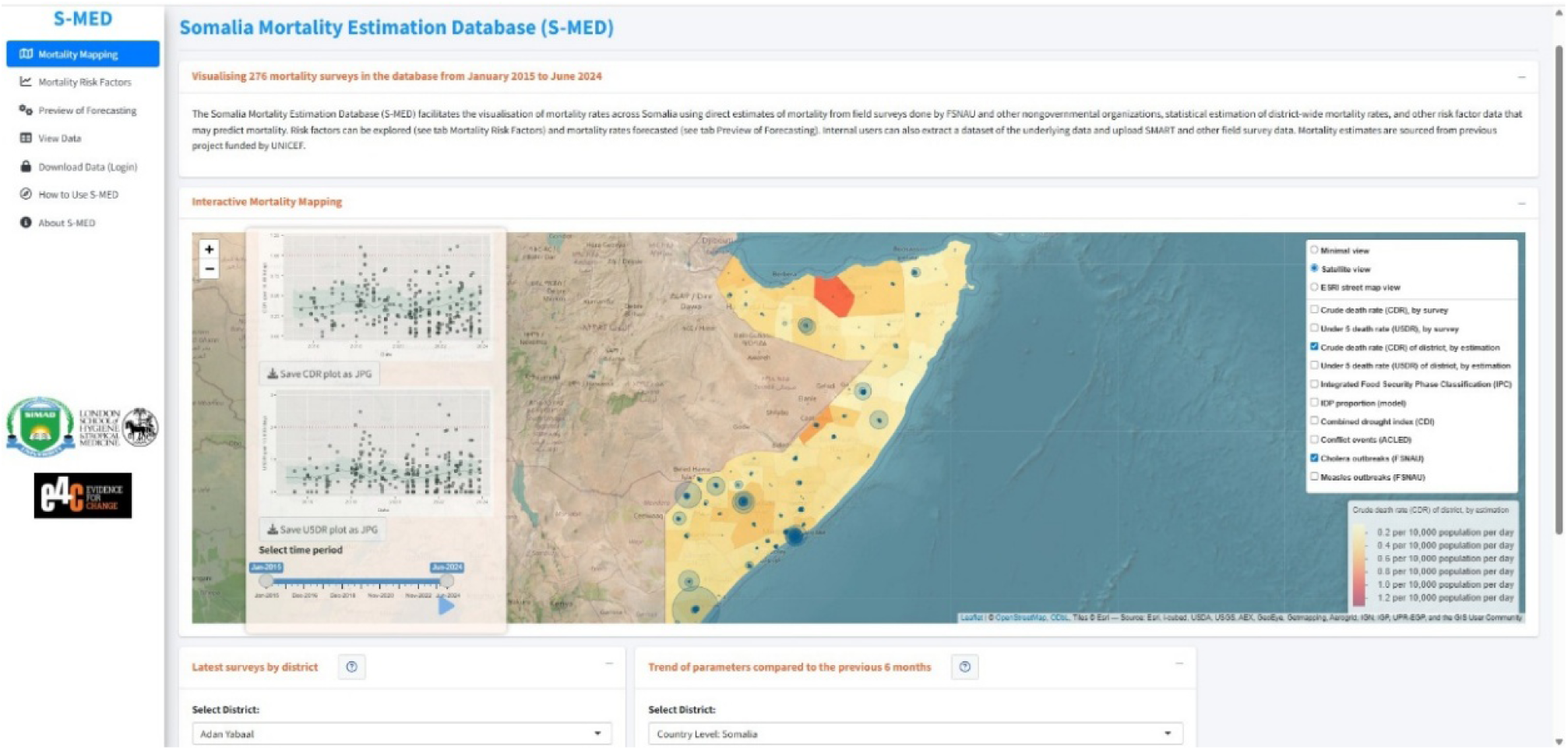
Frontpage of the Somalia Mortality Estimation Database (S-MED)

### Study population, period, and outcomes

We included all populations affected by crises in Somalia including pastoral, agropastoral, riverine, and urban populations; displaced and resident populations; and populations across all Federal Member States and regional administrations including Somaliland and Puntland.

Survey-based estimates were stratified at the district level (mean population 200,000, 74 districts total) and by month. The main outcomes were CDR and U5DR, the number of all-age and under-5-year deaths per 10,000 person-days, respectively.

### Data sources

All data sources are in the public domain, except for SMART surveys and disease incidence, which were shared by UNICEF under a previous aligned project on mortality estimation (UNICEF ESARO, grant number 2022/1291432-0) (see Table 1). The 10-year period covered January 2015 to June 2024 inclusive, to coincide with the data available to us. Mortality estimation based on modelling was sourced from a previous project.[14]

**Table 1.** Predictor data used in the Somalia Mortality Estimation Database (S-MED)

| Potential predictor | Justification | Index | Source |
| --- | --- | --- | --- |
| Integrated Food Security Phase Classification (IPC) | Indicator of stressed food security to famine conditions | (1) Minimal/none<br>(2) Stressed<br>(3) Crisis<br>(4) Emergency<br>(5) Catastrophe/famine | Integrated Food Security Phase Classification (IPC) (https://www.ipcinfo.org/), via R package ripc (https://github.com/ocha-dap/ripc) |
| Proportion of IDPs among total population | Forced displacement predicts crude mortality due to lack of protection, poor access to curative and preventative healthcare, food security and WASH | 0 to 100% | International Organization for Migration (IOM) Displacement Tracking Matrix (https://dtm.iom.int/), via R package dtmapi (https://displacement-tracking-matrix.github.io/dtmapi-R/) |
| Violent events | Violent events signify protection risks and reduced access to services and markets | Count of events | Armed Conflict Location and Event Data (ACLED, https://acleddata.com/) via R package acled.api (https://gitlab.com/chris-dworschak/acled.api) |
| Suspected and confirmed cholera cases | Epidemic disease burden | Count of cases | Food Security Nutrition and Analysis Unit (FSNAU, https://fsnau.org/) |
| Suspected and confirmed measles cases | Epidemic disease burden | Count of cases | Food Security Nutrition and Analysis Unit (FSNAU) |
| Combined drought index (CDI) | CDI combined rainfall, temperature and a proxy of soil moisture to assess susceptibility to drought (drought impacts livelihoods, food security, and access to WASH) | (1) >1.0, no drought<br>(2) 0.8—1.0, minimal drought<br>(3) 0.6—0.8, moderate drought<br>(4) 0.4—0.6, severe drought<br>(5) <0.4, extreme drought | Somalia Water and Land Information Management (SWALIM, https://cdi.faoswalim.org/index/cdi) |

### Ground data on mortality from small areas

Data sources included retrospective surveys (where households are interviewed about all demographic events over a recall period to produce mortality estimates and confidence intervals, across populations at the district-level or in camps) and prospective surveillance (typically small-scale vital registration systems where regular household visits by community workers provide information on demographic events and changes to the population).[4] The retrospective household surveys used the SMART methodology which combines estimation of acute malnutrition, CDR, and U5DR.[3] Interviewers solicit information from respondents on the demographic evolution (composition, births, deaths, in- and out-migration) of their households during a recall period of three to four months. They use systematic random sampling or two-stage cluster sampling with probability of cluster selection proportional to size and typically target district, or urban areas or internally displaced person (IDP) settlements within a district. Repeated, monthly surveillance estimates of CDR and U5DR from 57 IDP camps across Deynile and Kahda districts, Banadir region, from April 2016 to November 2023 were also integrated.[15] Estimates of U5DR from the Nutritional and Mortality Surveillance System in IDP camps in Afgooye (from July 2022) were added [16, 17]. This surveillance systems produce population-based estimates wherein community health workers doing monthly household visits update nutrition, mortality, and vaccination status and conduct simplified verbal autopsies.

### Modelled data on mortality at district level

To improve the geographical coverage and estimate mortality at the district level, statistically modelled estimates from previous analysis done by LSHTM and SIMAD were included [14]. A series of candidate statistical models, each combining different district-level predictors, were trained and validated using the SMART survey dataset. Model performance was assessed through leave-one-out cross-validation (LOOCV) on held-out observations. The final model was selected based on the best root mean square error (RMSE) and used to estimate mortality under observed conditions, producing district-level estimates of the CDR and U5DR. Full methodological details are described in Ouchtar et al.[14]

### Predictor data on mortality at district level

Data on potential predictors of mortality by time were included to visualize and analyze alongside CDR and U5DR, in accordance with previous analyses (see Table 1) [10, 14].

### Analysis

#### Spatiotemporal visualization and risk factor analysis

On the front page of the dashboard (Figure 2), the user can visualize CDR and U5DR and their 95% confidence intervals (CIs) by moving the time slider over the period of interest to show spatiotemporal changes in mortality over time. This leads to (a) the plotting of a trend line in the inset panel for CDR and U5DR against the commonly used Sphere-endorsed crisis thresholds of 1 death and 2 deaths per 10,000 person-days, respectively, and (b) the display of mortality estimates and selected predictor variables on the map of Somalia, by level of intensity.[18] The bottom panels provide survey estimates and comparisons of data during the last six months, for the selected district or across Somalia. On the second tab, predictor variables and modelled district-level CDR and U5DR and their 95% CIs are plotted by district (or nationally) to display the potential correlation with the proportion of IDPs, combined drought index (CDI), violent events, severe outbreaks, and mortality. This can be used to explore hypotheses of correlation between predictors of mortality and their estimates, at the district level. For instance, as Somalia has two annual rainy seasons (the Gu and Deyr rains, expected during April to June and October to December, respectively), the onset of severe drought in the dry season may be correlated with severe food insecurity, famine-associated disease outbreaks, malnutrition and ultimately increased CDR and U5DR after a lagged period of several months. Localized outbreaks of cholera may cause more rapid-onset spikes in mortality in affected districts. An influx of IDPs may cause pressure on scarce health and food resources, thus gradually increasing population-level mortality, with IDPs themselves at high risk of death owing to a lack of health and food resources, and endemic and epidemic disease.

#### Case studies of the use of S-MED for visualization of deteriorations in health status

To show how S-MED can be used, we investigated whether signals of early deterioration in health status would appear before a rise in mortality, through a delayed impact on CDR and/or U5DR, and whether these incidents could be detected in different locations in Somalia. We examined the cases of the food security crisis in 2017—2018 [10] and drought-induced mortality fluxes in 2022 to mid-2023 [14]. By aggregating mortality data, S-MED can facilitate more complex analyses including crisis-wide estimation of population mortality[19], geospatial analysis[20], and forecasting of mortality rates.[14] These analyses will not be discussed in this paper.

#### Ethical review

This study was approved by the Research Ethics Committee of the London School of Hygiene and Tropical Medicine (#31136) and SIMAD University, Somalia (#2024/SU-IRB/FMHS/P0012).

## Results

We compiled CDR and U5DR from 276 survey-based estimates from January 2015 to June 2024 and from 28 (37.8%) of the 74 districts, and surveillance-based estimates from community-based surveillance systems in IDP camps in Mogadishu (October 2022 to November 2023) and Afgooye (July 2022).[15, 16] More than 80% of the estimates came from south and central regions (48%) including Banadir (n=31), Bay (n=30), Hiran (n=25), Lower Juba (n=23) and Mudug (n=20); from Puntland (20%) including Bari (n=28) and Nugaal (n=25) regions; and from Somaliland (14%) including Togdheer (n=16) and Woqooyi Galbeed (n=22) regions. Using the CDR and U5DR emergency thresholds of 1 and 2 deaths per 10,000 person-days respectively, above-threshold estimates were found primarily in Bay and Banadir in 2022 and Nugaal in 2018. Focusing on urban IDPs, surveillance in the Mogadishu IDP camps reported CDR (1.26 to 3.15 deaths per 10,000 person-days) and U5DR (2.7 to 7.05 deaths per 10,000 person-days) more than three times the threshold in October 2022 to February 2023. [16] Visualization of datasets can give insights into potential determinants of mortality. Figure 3 shows the convergence of increased U5DR in south central areas including Mogadishu, Baidoa, and Lower Juba (in yellow and orange shading) and its confluence with conflict events (i.e. political violence, explosions, remote violence, and Al-Shabaab violence targeting civilians) (green circles) in 2023 to mid-2024. This corresponds with an analysis by ACLED on the concentration of attacks targeting civilians in Lower Juba and Bay, and of Al-Shabaab-driven violence in Lower Shabelle, where African Union Transition Mission bases are located.[21]

**Figure 3.**
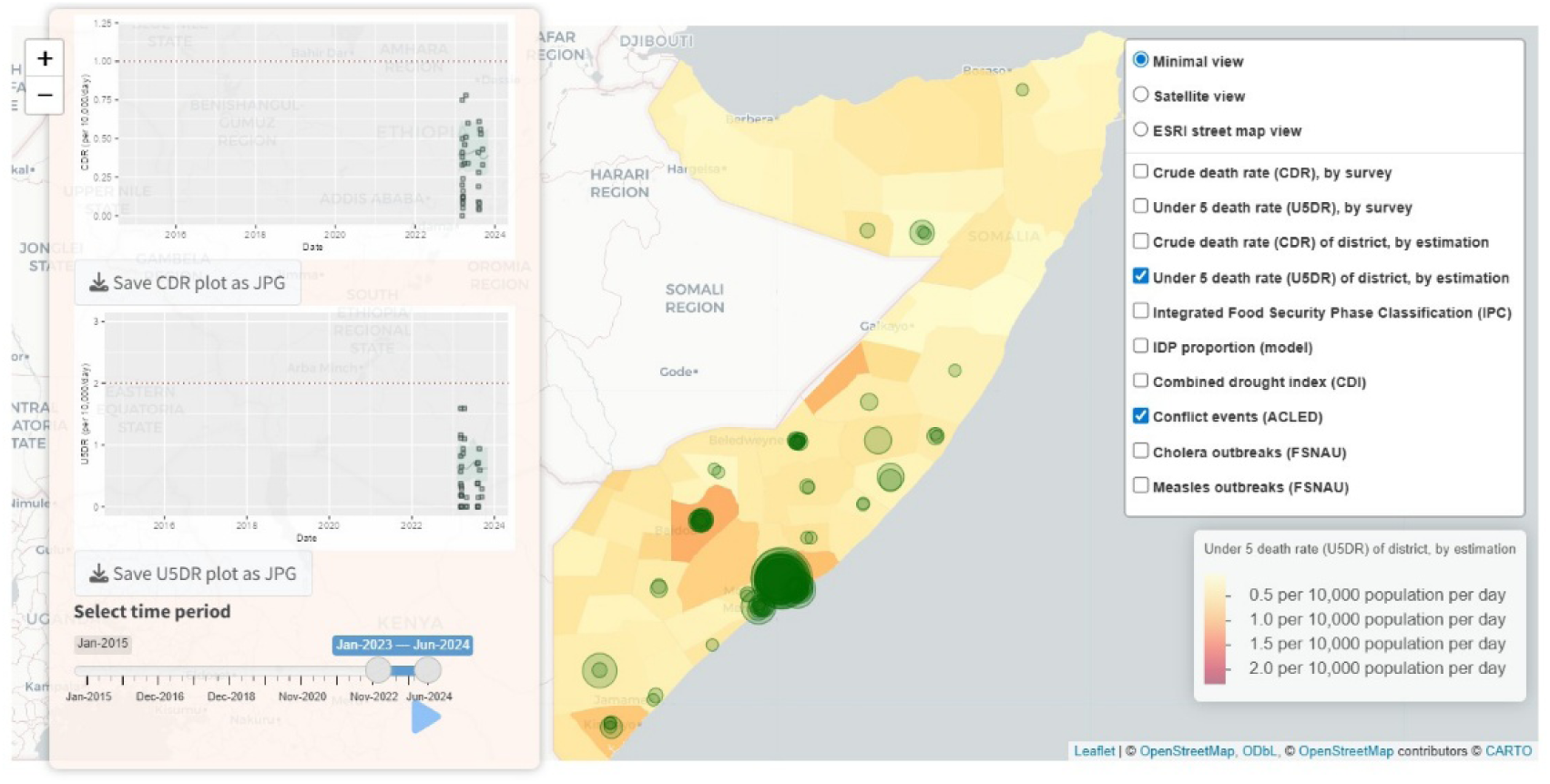
Convergence of modelled under 5 death rates (yellow to red sharing) and conflict events targeting civilians involving Al-Shabaab and security forces (green circles) in 2023 to 2024

### Case study 1: Crisis indicators for drought-associated mortality during the 2017 and 2018 food security crisis

From September to December 2016, continuous, poor seasonal rainfall befell most regions of Somalia.[22] This severely hampered the harvests and livestock survival, and by early 2017 ushered in drought, large-scale distress migration (often from urban to rural areas due to the search for food and water), and famine- and displacement-associated epidemics of measles and cholera. Conflict-driven displacement was seen in Lower and Middle Shabelle and Banadir (in the south) and Sool, and Toghdeer (in Somaliland).[10] The health and nutrition response was weakened due to chronic lack of investment in the health system, poor vaccination coverage, and generally poor humanitarian access in the midst of Al-Shabaab-driven violence.[9] IPC Phase 4 (emergency) conditions were projected from February to June 2017 for districts in southern regions and large parts of Puntland, with Somaliland remaining in IPC phase 3 (serious) [23]. From January 2017 to mid-2018, a large-scale loss of life, measured as excess CDR up to 0.09 per 10,000 persons-days higher than the most likely counterfactual CDR, was estimated across Somalia.[10] This occurred at a large geographical scale, across areas classified as relatively modest IPC Phase 3 (crisis). By mid-2018, there was above-average rainfall, and a decline in CDR was observed.[10]

### Available tools and their use for situational analysis

Nutrition surveys conducted by FSNAU and partners in late 2016 showed critical levels of acute malnutrition in half of assessed populations, particularly among IDPs. Critical-level global acute malnutrition (GAM) prevalence (≥15%) was reported in half (13/27) and very critical severe acute malnutrition (SAM) (≥4%) in nearly a quarter (22% or 6/27) of the nutrition surveys conducted in rural and IDP communities, indicating an increase in malnutrition prevalence compared to July 2016. Famine early warning systems including the FSNAU dashboard and FEWS NET tracked predictor data on rainfall, market prices, food access, and nutrition survey results to project food security outcomes and potential food security scenarios.[24] Conflict and outbreaks of measles and cholera co-occurred with displacement, particularly in IDP-concentrated urban centres (Mogadishu, Baidoa, Belet Weyne). Survey-based estimates later revealed CDR and U5DR levels well above emergency thresholds throughout 2017 and 2018.[10] Among IDPs in the Afgooye corridor, U5DR reached 7 deaths per 10,000 person-days in 2017.[16] The estimated excess death toll in 2017 rose to double that of 2016, with the highest excess CDR in the northeastern regions and Hiraan and highest excess U5DR in southcentral regions.[10] Mortality surveillance typically exists in a void wherein it is not compiled or incorporated into weekly disease surveillance reporting by the Early Warning Alert and Response Network (EWARN) or Integrated Disease Surveillance and Response (IDSR).

While the dashboard data and its visibility narrowed the gap between early warning and action, it did not make a link directly with health, nutrition, and mortality outcomes or use a narrative to provide a situational analysis.[24] This made it difficult to understand the meaning of predictive data, and the progressive impact on at-risk populations via mortality and malnutrition assessments.[24]

### Use of S-MED for further situational analysis

By April 2017, conditions in parts of southcentral regions and Puntland met the criteria for extreme drought using the CDI (specifically, CDI < 0.4; i.e., “major loss of crops and pasture, extreme fire danger, total water shortages, drying of deep reservoirs and usage restrictions”). However, this signal did not appear to be linked as an early warning of morbidity and mortality risk at the time. With S-MED, users could have visualized the deteriorating health situation with more clarity. The early CDI signal showing widespread CDI at the moderate level (0.6—0.8) was observable throughout Somalia prior to the Gu rainy season and as early as May 2016, particularly in southern and northeastern regions, and CDI at the extreme level (<0.4) by November 2016 after repeated failures of the rain is clear in Figure 4A. This can be viewed alongside intensifying displacement towards rural areas, growing cholera and measles outbreaks in southcentral urban areas that had a high concentration of IDPs, indicating sub-optimal measles vaccination coverage, overcrowding, and exposure to unsafe water and sanitation (Figure 4B). Distress migration started in November 2016, with IDPs moving closer to food and water [9].

**Figure 4A-C.**
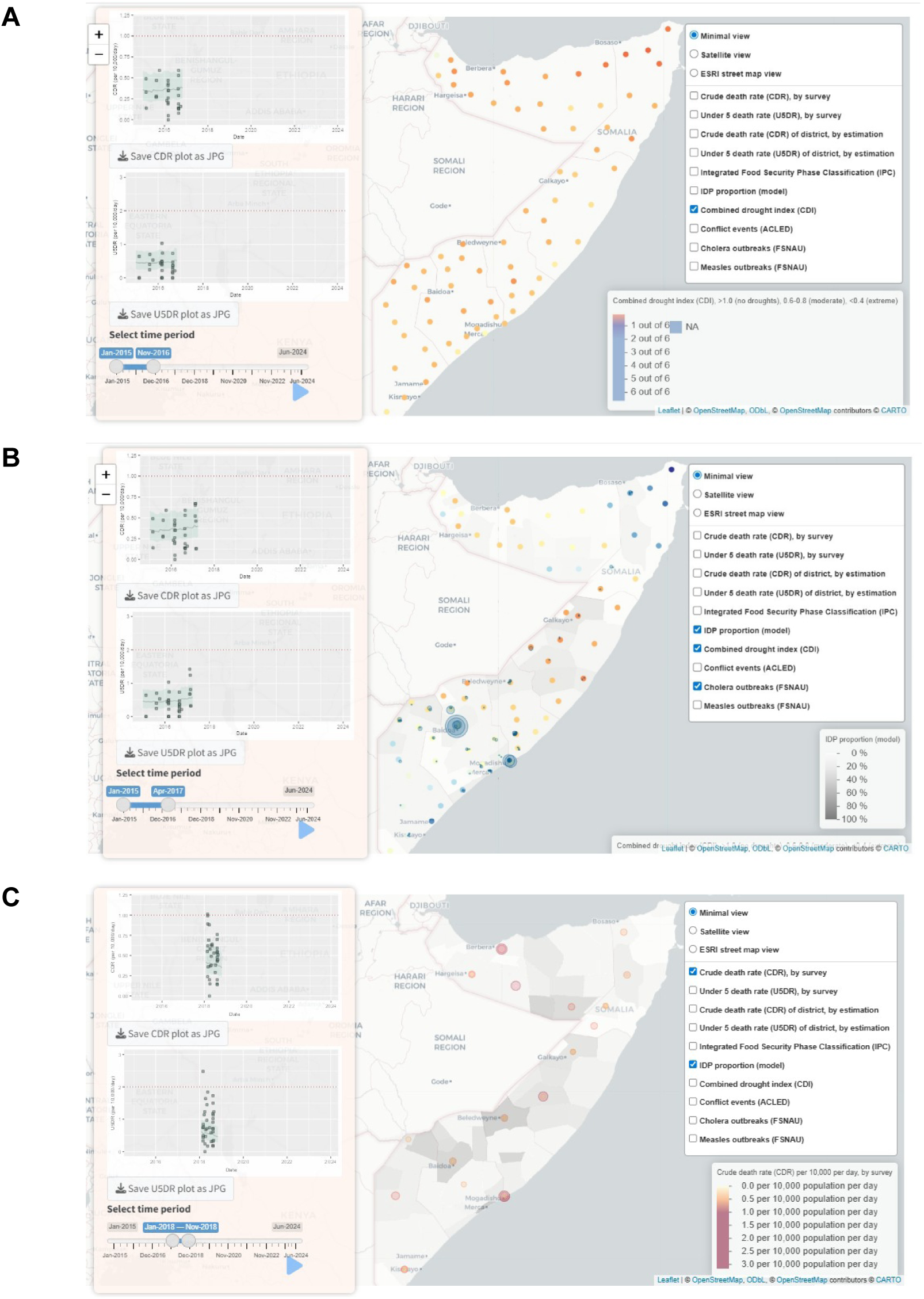
(A) Early signal from combined drought index (CDI) at extreme levels across Somalia in Nov 2016 (red dots); (B) cholera outbreaks (dark blue) in Southcentral Somalia combined with extreme CDI in Apr 2017; (C) hotspots of high crude mortality (trend lines beyond threshold and orange and red spheres) and measles outbreaks (red dots) in high-displacement locations in Southcentral, Puntland, and Somaliland, Nov 2018.

The combined presence of drought, epidemic disease, malnutrition, and forced displacement predated above-threshold CDR and U5DR measured by surveys in early 2018 in Mogadishu, Somaliland, and Puntland (Figure 4C). The detected spatial convergence of conflict, drought, and epidemics in southcentral regions could have potentially triggered earlier actions to prevent the loss of life. Estimates of U5DR among IDPs in 25 camps in the Afgooye corridor outside Mogadishu reached 7d/10000/day, primarily among new arrivals during an influx of IDPs into the settlement in May and June 2017 [16]. The main causes of death, by verbal autopsy, were diarrhoeal diseases, measles, and SAM, illustrating the confluence of preventable disease and food insecurity seen in S-MED data streams. From April 2018 onwards, increased rainfall during the Gu season improved food security, market conditions and harvest; by the first quarter of 2019, CDI increased in southcentral regions and Puntland, few outbreaks were reported, and CDR and U5DR remained below alert thresholds throughout 2019.

### Case study 2: Drought-induced mortality surge in South Central, 2022—2023

By late 2021, early warning signals for drought were issued following the failure of the second consecutive Deyr rainy season (October to December), and widespread drought, poor harvests, and livestock deaths took hold by early 2022. These poor livelihood conditions were exacerbated by violence and ethnic tensions, massive internal displacement, increasing global food prices, and socioeconomic repercussions of the COVID-19 pandemic. Throughout 2022, retrospective analysis found that 43,000 excess deaths occurred, with half occurring among children under 5 years.[25] As compared with the 2017—2018 period, most of the deaths were concentrated in the south and central regions without Puntland. The highest CDR and U5DR were measured in Bay, Bakool, and Banadir (with its large IDP population, in particular).[25]

### Available tools and their use for situational analysis

IPC Phase 4 (emergency) conditions were projected in southcentral regions in late 2021, and FEWS NET, SWALIM, and the FSNAU dashboard all issued alerts on the impending drought based largely on environmental and food security predictor data.[26] Mortality estimates were established in multiple sites in southcentral regions in May, which indicated near-threshold conditions (CDR = 0.75 to 0.86 deaths per 10,000 person-days; U5DR up to 1.88 deaths per 10,000 person-days) had been reached in late January. By July 2022, the Building Resilient Communities in Somalia (BRCiS) Consortium implemented mortality surveillance in 11 IDP settlements with the aim of providing real-time information on the impending crisis and state of the humanitarian response.[17] Over a 1-month recall period in 2022, CDR was recorded at 0.94 deaths per 10,000 person-days and U5DR at 3.03 deaths per 10,000 person-days, exceeding emergency thresholds, and largely due to recent influxes into the camps and the confluence of measles, cholera, acute malnutrition and inadequate WASH, malnutrition, and vaccination coverage.[17] This concorded with the IPC Phase 4 classification. By October 2022, most regions were classified as IPC Phase 4 (emergency) and two southwestern regions were classified as IPC Phase 5 (famine). Mortality estimates were still derived from retrospective surveys conducted by humanitarian agencies, typically months after the mortality peaks. For example, high mortality rates were documented among IDPs in Baidoa and Mogadishu via surveys conducted in late 2022, but indications of deteriorating conditions occurred as early as December 2021. These retrospective findings, including mortality rates exceeding emergency thresholds (e.g., 3.15 deaths per 10,000 person-days in Mogadishu IDP camps), confirmed the severity of the crisis but did not inform real-time prioritization of response. Consequently, despite improved technical tools and more robust humanitarian coordination, prompt action remained limited by the lack of mortality data integrated into situational analyses.

### Use of S-MED for further situational analysis

If S-MED had been operational in real time, it could have been used to identify hotspots for potential increases in excess mortality, by tracking CDI decline (with the early signal in southcentral regions from November 2021 shown in Figure 5A and deterioration shown in Figure 5B), alongside early indications in January and February 2022 of increased incidence of conflict events and displacement, and near-threshold CDR and U5DR in IDP camps in southcentral regions (Figure 5C). By October 2022, S-MED was able to show the overlap between famine declaration as per IPC Phase 4 (emergency) across Somalia and above threshold CDR and U5DR in the map and the adjacent CDR and U5DR time series (Figure 5C). Use of the risk factor analysis illustrates in Galgaduud for example, after the initial drop of CDI <0.4 in January 2022, an apparent increase in CDR and U5DR toward near-threshold levels followed in the next three months (March 2022) and was sustained until August 2022 (Figure 6).

**Figure 5A-C.**
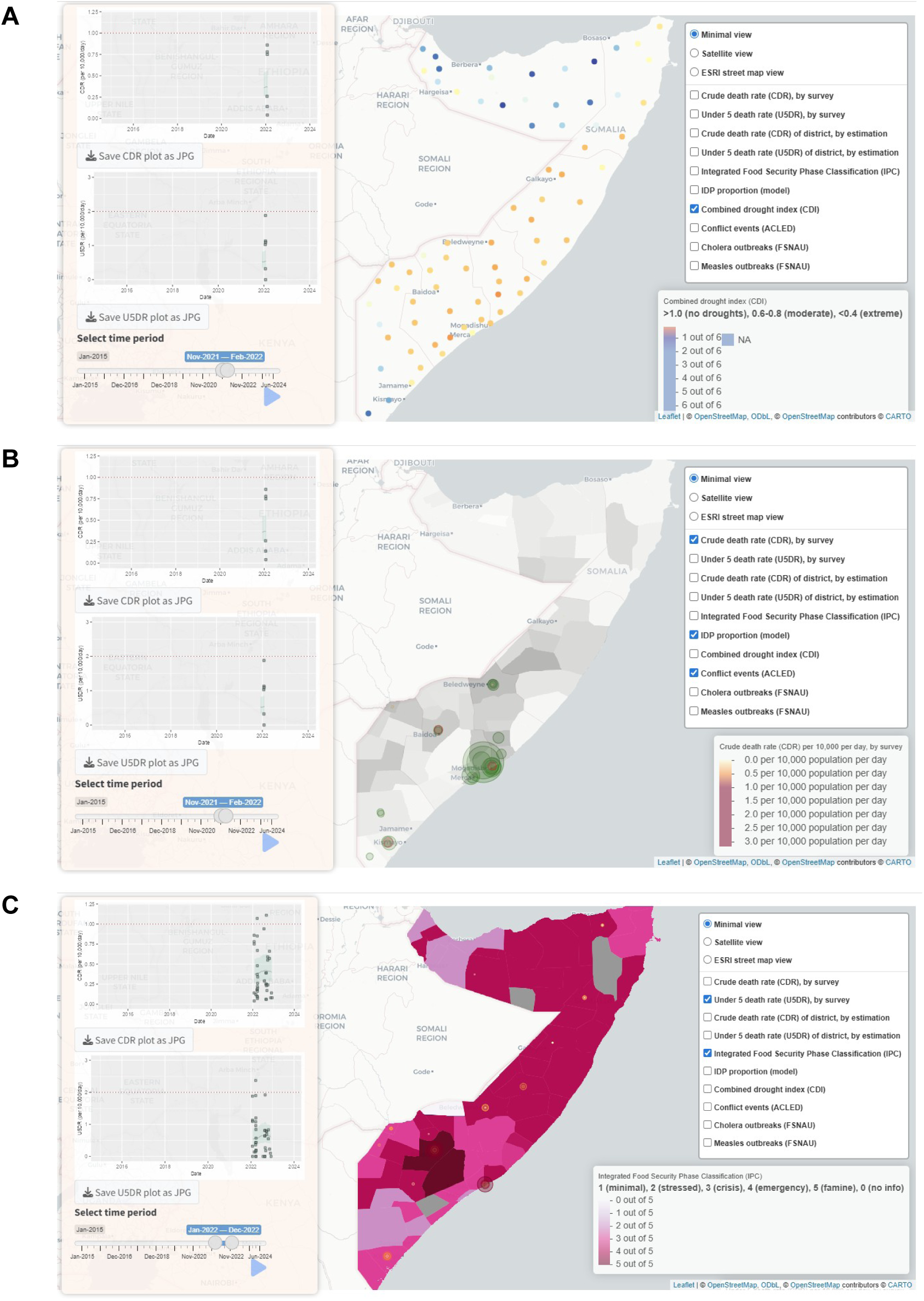
(A) Early signal from combined drought index (CDI) at extreme levels in southcentral Somalia, in Nov 2021 (orange, red dots); (B) conflict events (green circles), displacement (grey shading) and extreme CDI (orange, red dots) in southcentral Somalia by Feb-Jun 2022; (C) integrated food security phase classification (IPC) of 4 (emergency) and 5 (famine) by Dec 2022 coinciding with above-threshold CDR and U5DR.

**Figure 6.**
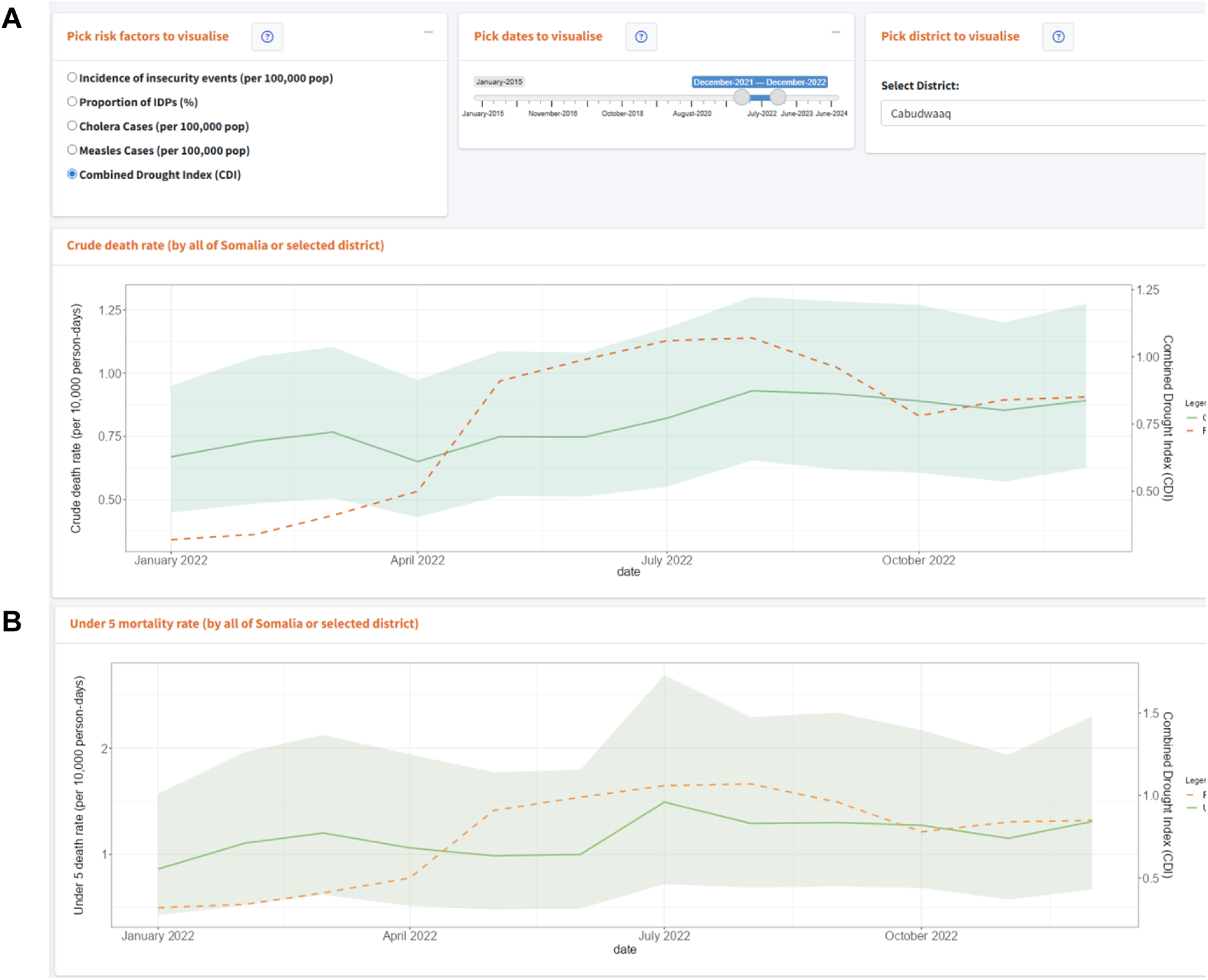
Risk factor analysis of combined drought index (CDI) as predictor of estimated crude and under-5 mortality in Cabudwaaq, Galgaduud, Somalia, December 2021 to December 2022. Note that CDI < 0.4 indicates extreme drought; 0.4—0.6, severe drought, and 0.6—0.8, moderate drought.

## Discussion

Communities across Somalia are routinely subjected to a cascade of intersecting seasonal hazards (including poor rainfall and drought, loss of livelihoods, forced displacement due to the search for food and water, crowding in informal settlements, acute malnutrition, coincident epidemics, and disrupted health systems) and Al-Shabaab-driven conflict. We used S-MED to explore case studies of emerging crises in Somalia from 2015 to 2024 by positing that if the integration and visualisation of morbidity, environmental, and mortality data seen in the repository were collated and visualized in real-time in S-MED, we could provide a bird’s eye view of mortality and the predictive value of morbidity and environmental risk factors across space and time. Public health interpretation can be used to generate insight into early signals of risk factors that in a causal chain, lead to excess mortality.[12] Insight from excess mortality rates, while a late crisis indicator that demonstrates a crisis in progress, can be used for improved public health situational analysis, alongside famine projections and scenario-building via the well-established IPC and FEWS NET, respectively. Essentially, capturing mortality trends and sharp increases in short time periods, within the complex framework of food insecurity and malnutrition projections, would provide empirical data to reinforce the typical triggers for urgent humanitarian action for vulnerable populations. Other risk factors, such as CDI, could be used even earlier and in unison with emerging mortality signals to show the progression of a severe drought and its health impacts, at the earliest stages. Further combination of monthly, district-level food insecurity early warning datasets (e.g., the Harmonized Food Insecurity Dataset) with ad-hoc mortality estimates, district-wide mortality estimation and armed conflict events could enable the systems-level thinking to drive comprehensive, multisectoral analysis of drought, food insecurity, and morbidity and mortality outcomes, and predictive modeling.[27]

S-MED provides a digital platform to compile these data, but it is truly a human-centered process that critically depends on relationships between national and international organizations and governments who collect and need to share mortality data, and the judgement of humanitarian analysts who understand the context and who can make sense of diverse and shaky data and their relationship with malnutrition, severe morbidity, and mortality. Maxwell and Hailey (2020), in their analysis of the potential and pitfalls for famine early warning systems and early action in East Africa, stated the obvious but elusive necessity of NGOs and governmental actors, as well as information actors and programmatic actors to work together to share, interpret, and act on data and be accountable to the program cycle that links early warning with early action.[24] These tenets ring true for a mortality data repository. First, to provide a true picture of mortality across time and space, there is a need to work closely with NGOs and other agencies who choose to share their mortality estimates via survey reports and datasets on a timely basis, driven by a common perceived benefit to better understand the broader picture of mortality and its determinants, and a common good in data sharing for accountability to populations. This could be facilitated through the Health and Nutrition Clusters’ use of S-MED to coordinate future health, mortality, and nutrition assessments by highlighting areas of poor coverage, presumed deterioration, and influxes of displacement.[5] We hope that it will highlight areas that are consistently left out of mortality estimation, including areas of high violence, high population density, and the resulting lack of humanitarian access.

Generating hypotheses and building out scenarios based on these data is often challenging and requires proficiency in understanding causal and temporal relationships between upstream indicators (i.e., conflict events, CDI), downstream indicators (i.e. epidemic incidence), and impacts (i.e., malnutrition and mortality). For instance, linking a high U5DR from retrospective surveys with a declining and lower-than-expected current prevalence of SAM during a food crisis and coincident measles epidemics requires investigating whether malnourished children have already died. This can be translated into ‘alarms’ to signal a deteriorating situation, though this risks masking causal mechanisms and identification of specific humanitarian needs (i.e., a widespread lack of inpatient malnutrition treatment). Less is known about linking early warning signals for famine with the evolving malnutrition and mortality situation demonstrated through field studies. In a retrospective meta-analysis of hundreds of mortality surveys in Somalia, Warsame and colleagues found that among populations with a moderate IPC grading of 3 (crisis), excess mortality accumulated over time, highlighting the cascade of other risk factors that should be considered for true early warning that could spur early action. Across East Africa, the question of how to most efficiently link well-identified and valid early warning signals and analyses on famine, mortality, and malnutrition with a highly-sensitive policy and humanitarian response continues to be an area of focus.[24] In addition, more fine-scale detail linking mortality estimates with livelihood groups and clans that may be particularly vulnerable to hazards is needed.

### Next steps for S-MED

We plan to solicit feedback on this iteration of S-MED from key actors in the Somalia humanitarian ecosystem, with a view to the major USAID-driven humanitarian funding cuts that have taken place in 2025, and the need for more collation of mortality and malnutrition assessments and more efficient situational analysis. We aim to liaise with a major consortium of NGOs who conduct sub-district level mortality surveys and community-based organizations operating community-based surveillance systems. This may increase the yield of estimates from more silent, poorly accessed, and volatile areas. Data quality indicators, first established by the CEDAT project, can be used to assess the trustworthiness of survey and surveillance design, sampling, and reporting to potentially flag highly-biased or non-representative reports.[7] We will solicit for more data on key determinants of mortality, prioritizing data on the prevalence of global and severe acute malnutrition (from SMART nutrition surveys), vaccination coverage (from UNICEF-funded vaccination coverage surveys), availability of health services (i.e., Health Resources and Services Availability Monitoring System (HeRAMS), as available), terms of trade purchasing power index to better understand changes to food insecurity and livelihood[10], climate data used for anticipatory action in humanitarian assistance (i.e., daily precipitation level and temperature), and a mapping of the presence of humanitarian actors.[10] With the accumulation of surveys and surveillance data, we can plan wide-area analyses of seasonal and episodic increases in excess mortality, to investigate the magnitude, and the major determinants amongst the predictor variables and their relationship to early warning signals. We aim to validate and include a predictive forecasting method to project mortality rates likely to occur over the next one to three months for small areas and populations where there is sufficient data. This will help to add more value to the retrospective data and could serve as an additional early warning signal. Finally, we will adapt the application to a mobile-friendly version.

As a digital platform, S-MED is used to compile the work of several public health actors and is therefore only as useful as the real-world collaboration amongst those actors, and in turn the feedback that the analyses give them to improve their work. S-MED provides a framework to apply to other countries in crisis that require collation and interpretation of small-scale mortality estimation done across its territory. We encourage use (and improvement) of the framework and code to better characterize, explain, and predict crude and under-five mortality across humanitarian settings.

## Data availability

S-MED can be accessed at https://hhc-lshtm.shinyapps.io/S-MED/. The code and underlying data for S-MED is publicly available at https://github.com/ruwanepi/S_MED.

